## Supplementary materials for "Using patient biomarker time series to determine mortality risk in hospitalised COVID-19 patients: a comparative analysis across two New York hospitals"

June 29, 2022

### 1 Supplementary materials

#### 1.1 Biomarker abbreviations

Throughout this article, we use standard abbreviations for biomarkers: the full names corresponding to these acroynms are given in Table S1.

#### 1.2 Data processing

##### 1.2.1 Maimonides data processing

We now detail the steps taken to clean and process the data to make it into a form amenable to estimation. We obtained demographic and patient history for $n = 999$ individuals who were admitted to Maimonides hospital. A number of patients ($n = 11$) were admitted to hospital well before what we deemed the start of the pandemic (which we took as 2nd February 2020). Since these patients were likely admitted for different conditions and contracted COVID-19 in hospital, they were removed from the analysis. There was $n = 1$ individual whose demographic data could not be matched with the test data, and they were removed from the dataset. This resulted in $n = 987$ patients that whose demographics could be matched with their outcome.

**Initial data processing for dynamic analyses** For the analyses involving time-dependent measurements, we carried out an additional series of transformations on the data. A number of observations ($n = 518$ observations corresponding to $n = 14$ patients) were recorded as occurring before a patient was recorded as admitted: for these patients, we changed their date of admission to be the date when their earliest observation was recorded. There were $n = 1474$

| Full name | Abbreviation |
|---|---|
| Alkaline Phosphatase | ALKP |
| Basophil Percentage | BASO PCT |
| Brain Natriuretic Peptide | BNP |
| Blood Urea Nitrogen Level | BUN |
| Serum C Reactive Protein Level | CRP |
| Eosinophils Percentage | EOS PCT |
| Blood Potassium Level | K Bld |
| Lactate Dehydrogenase | LDH |
| Absolute Large Unstained Cells Count | LUC ABS |
| Large Unstained Cells Percentage | LUC PCT |
| Absolute Lymphocyte Count | LYM ABS |
| Lymphocytes Percentage | LYM PCT |
| Mean Corpuscular Haemoglobin | MCH |
| Mean Corpuscular Haemoglobin Concentration | MCHC |
| Mean Corpuscular Volume | MCV |
| Absolute Monocyte Count | MONO ABS |
| Monocytes Percentage | MONO PCT |
| Mean Platelet Volume | MPV |
| Blood Sodium Level | Na Bld |
| Absolute Neutrophils Count | NEU ABS |
| Neutrophils Percentage | NEU PCT |
| Platelets Count | PLAT |
| Red Blood Cells | RBC |
| Red Cell Distribution Width | RDW |
| White Blood Cells | WBC |
| Chronic Obstructive Pulmonary Disease | COPD |
| Congestive Heart Failure | CHF |

Table S1: **Biomarker abbreviations.**

observations across $n = 176$ patients where the last observation was recorded after their recorded change of status date (i.e. when they either died or were discharged). Of these, there were $n = 53$ patients where their last recorded observation occurred more than 48 hours after their change of status date: for these, there was considerable uncertainty about their duration of stay in hospital, and they were removed from the analysis. There were $n = 574$ patients where there was a gap of less than one day between their last observation and their subsequent recorded change of status: in these cases, we updated their change of status date to that of the last recorded observation. There were also $n = 236$ patients where there was a gap of more than one day between the last observation and their subsequent change of status. For those $n = 165$ discharged patients within this group, we updated their change of status date to be that of their last recorded observation. The $n = 71$ expired patients within this group were removed from the analysis as this gap typically signalled that the patients or their relatives had opted for palliative care at this point. After removing those clinical tests where the machine / procedure was different between the two hospitals, we were left with $n = 279,086$ observations across $n = 863$ patients.

We then converted each patient's data into a regular day-block form: where, for each day the patient was in hospital, the patient had a single observation for each clinical test. To do this, we made a number of assumptions: if multiple tests of the same type were conducted on a patient on a given day, we took their mean as the daily observation; where a patient had a day when a specific test was not conducted, we assumed that day's test measurement was the same as on the last day it was conducted; when a patient was missing observations for a test until some days after their admission, we assumed the test measurements on these intervening days were the same as that from the first measurement. To reduce the impact of imputed observations, we included only those $n = 39$ tests in the analysis where at least 70% of the individuals had a test taken (at least once). There were $n = 737$ individuals who had been tested on at least 80% of the tests remaining after the previous step: these were the only individuals included in the Markov analyses which involved test observations; for the other Markov analyses involving only demographic or comorbidity variables, $n = 863$ individuals were included. Those remaining tests where more than 40% of observations were interpolated ($n = 15$ tests) were also removed. Additionally, the test for phosphorus levels was also removed since there were a substantial proportion of patients without any observations using this test. Overall, approximately 18% of dynamic test observations were imputed, meaning that 18% of patient-test-days had observations imputed from previous or subsequent days' observations for that patient a described above. Finally, any individuals where ethnicity was not recorded were dropped from the analysis. This meant that there were $n = 844$ patients included in the Markov analyses that did not use the dynamic test data; for those using the dynamic test data, $n = 721$ individuals across $n = 23$ were included (although some of the analyses limited the testing variables to the $n = 18$ tests in common between SUNY and Maimonides – this number excludes MCHC since it is calculated directly from

MCV and MCH).

**Relative change analysis.** For these analyses, we partitioned the influence of each clinical test into two separate variables: the initial measurement for each patient ("initial" meaning that the test was done within the first day of admission); and the percentage change in test value from this initial value. For those instances when an initial observation was zero, we set the percentage change as zero if all subsequent test values were also zero; alternatively, we calculated a percentage change versus half the first recorded non-zero value of the test.

**Absolute values analysis.** We conducted also a series of regressions using the absolute test values opposed to a combination of initial test values upon presentation at hospital and the relative dynamic values from these baseline measurements. The only processing step for the raw test data was to normalise the test data by subtracting the overall mean and dividing by the standard deviation.

#### 1.2.2 SUNY data processing

The approach used to process the patient data for SUNY followed closely to that for Maimonides. For a full description of the approach taken to clean the SUNY data, see the supplementary materials of (1).

### 1.3 Markov model code

```
data {
  int N; // num obs
  int patient[N]; // indicates patient identification
  int npatient; // num patients
  int state[N]; // either 1 (discharge), 2 (in hospital) or 3 (death) during each day
  int ncovs; // number of covariates included in regression model
  matrix[N, ncovs] X; // regressor matrix
 }
transformed data{
  vector[3] ones = to_vector(rep_array(1, 3));
  matrix[N, 3] mstate;
  for(i in 1:N)
    for(j in 1:3)
      mstate[i, j] = state[i] == j ? 1 : 0;
}
parameters{
  vector[npatient] a0_raw;
  real a0_top;
  real<lower=0> sigma_a0;
  vector[npatient] b0_raw;
```

```
  real b0_top;
  real<lower=0> sigma_b0;
  vector[ncovs] a1;
  vector[ncovs] b1;
}
transformed parameters {
  // non-centered parameterisation for intercept priors
  vector[npatient] b0 = b0_top + sigma_b0 * b0_raw;
  vector[npatient] a0 = a0_top + sigma_a0 * a0_raw;
}
model{
  matrix[N, 3] p;
  vector[N] psum;
  p[, 1] = exp(a0[patient] + X * a1);
  p[, 3] = exp(b0[patient] + X * b1);
  p[, 2] = to_vector(rep_array(1, N));
  psum = p * ones; // sums each row
p = p ./ rep_matrix(psum, 3); // normalises each row of probability matrix
  p = p .* mstate; // zeros any probs corresponding to states that weren't observed

target += log(p * ones); // likelihood

// priors
  b0_raw ~ normal(0, 1);
  b0_top ~ normal(0, 1);
  sigma_b0 ~ normal(0, 0.05);
  a0_raw ~ normal(0, 1);
  a0_top ~ normal(0, 1);
  sigma_a0 ~ normal(0, 0.05);
  b1 ~ double_exponential(0, 1);
  a1 ~ double_exponential(0, 1);
}
```

#### 1.4 Additional results

| **Parameter(s)** | **Prior(s)** | **Reasoning** |
|---|---|---|
| Discharge individual patient intercepts and related population-level summaries | $\alpha_{0i} \sim \text{normal}(\alpha_o^{\text{top}}, \sigma_\alpha)$, $\alpha_o^{\text{top}} \sim \text{normal}(0, 1)$, $\sigma_\alpha \sim \text{half-normal}(0, 0.05)$ | Allows individual variation if substantial evidence exists |
| Expiry individual patient intercepts and related population-level summaries | $\beta_{0i} \sim \text{normal}(\beta_o^{\text{top}}, \sigma_\beta)$, $\beta_o^{\text{top}} \sim \text{normal}(0, 1)$, $\sigma_\beta \sim \text{half-normal}(0, 0.05)$ | Allows individual variation if substantial evidence exists |
| Regression coefficients | $\boldsymbol{\alpha}_1 \sim \text{double-exponential}(0, 1)$ $\boldsymbol{\beta}_1 \sim \text{double-exponential}(0, 1)$ | Sparsity inducing |

Table S2: **Priors used to estimate Markov model.**

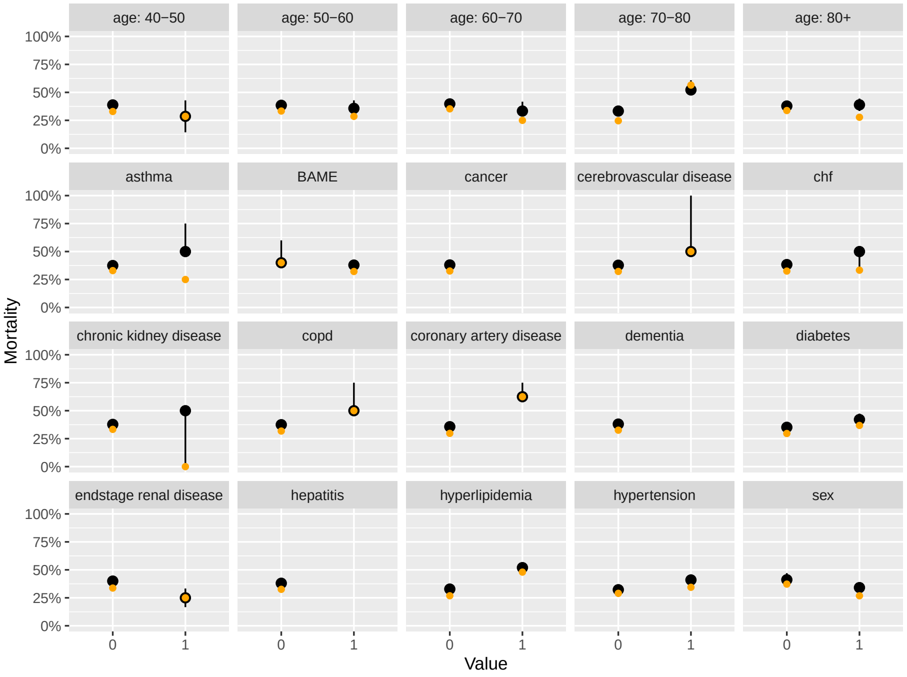


Figure S1: **Posterior predictive checks: demographic factors for SUNY predicting an independent SUNY dataset.** Each panel corresponds to a different binary variable, with the horizontal axis indicating its value. Orange points indicate observed mortality; black point-ranges indicate model estimated mortality, with the middle point indicating the posterior median, and the ranges indicating the 25%-75% ranges. The predictions were produced using the "post-admission" regression set in the Markov model.

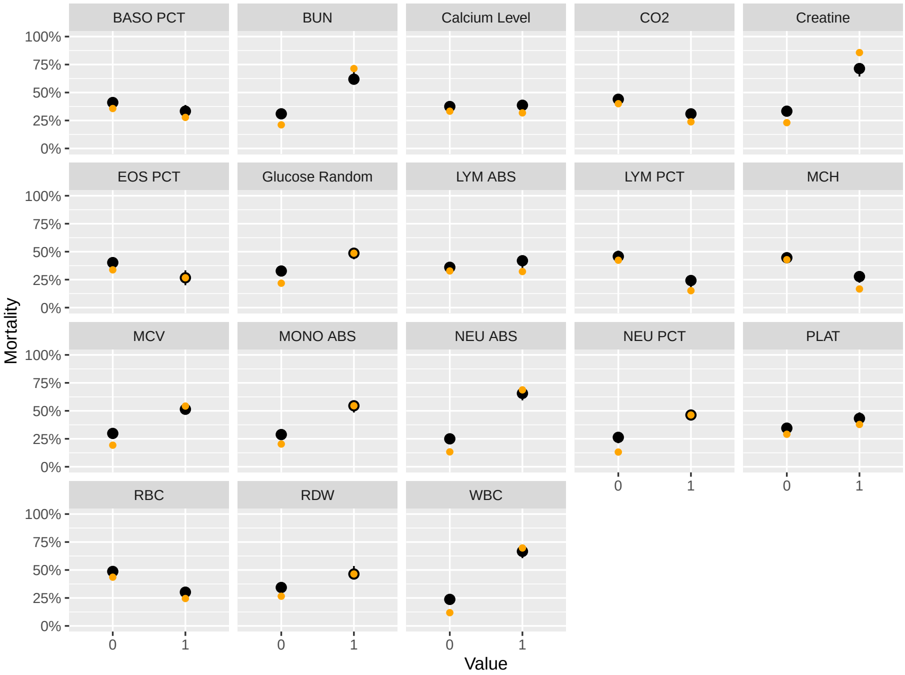


Figure S2: **Posterior predictive checks: dynamic factors for SUNY predicting an independent SUNY dataset.** Each panel corresponds to a different binarised variable (details of binarisation in text), with the horizontal axis indicating its value. Orange points indicate observed mortality; black point-ranges indicate model estimated mortality, with the middle point indicating the posterior median, and the ranges indicating the 25%-75% ranges. The predictions were produced using the "post-admission" regression set in the Markov model.

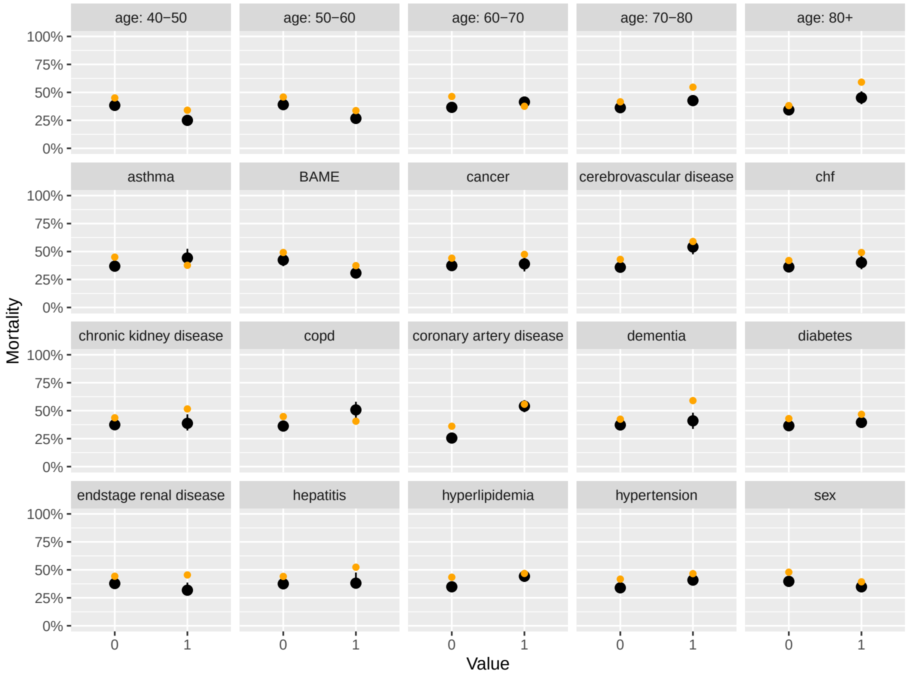


Figure S3: **Posterior predictive checks: demographic factors for SUNY predicting an independent Maimonides dataset.** Each panel corresponds to a different binary variable, with the horizontal axis indicating its value. Orange points indicate observed mortality; black point-ranges indicate model estimated mortality, with the middle point indicating the posterior median, and the ranges indicating the 25%-75% ranges. The predictions were produced using the "post-admission" regression set in the Markov model.

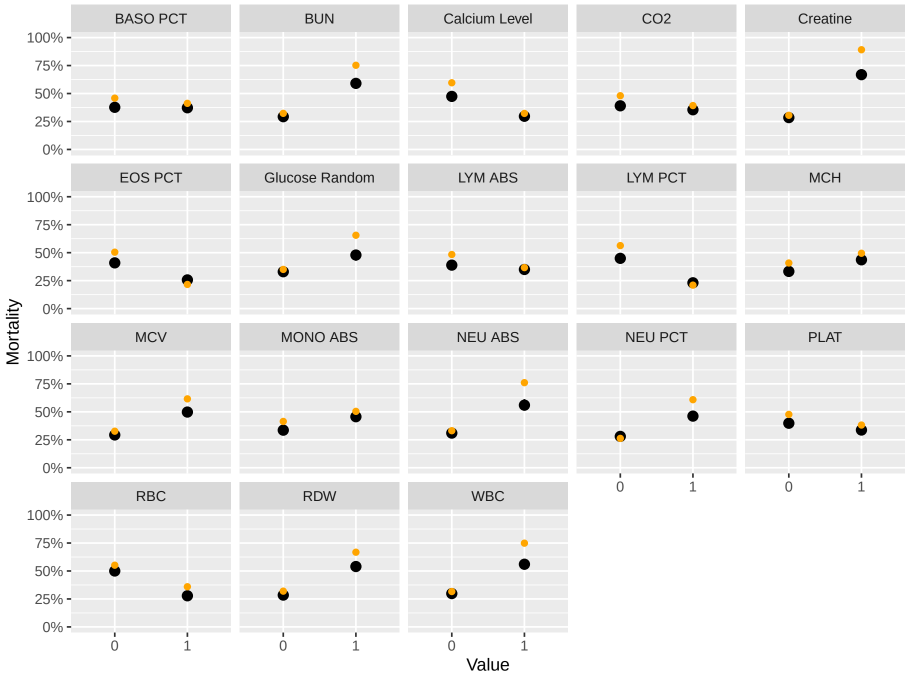


Figure S4: **Posterior predictive checks: dynamic factors for SUNY predicting an independent Maimonides dataset.** Each panel corresponds to a different binarised variable (details of binarisation in text), with the horizontal axis indicating its value. Orange points indicate observed mortality; black point-ranges indicate model estimated mortality, with the middle point indicating the posterior median, and the ranges indicating the 25%-75% ranges. The predictions were produced using the "post-admission" regression set in the Markov model.

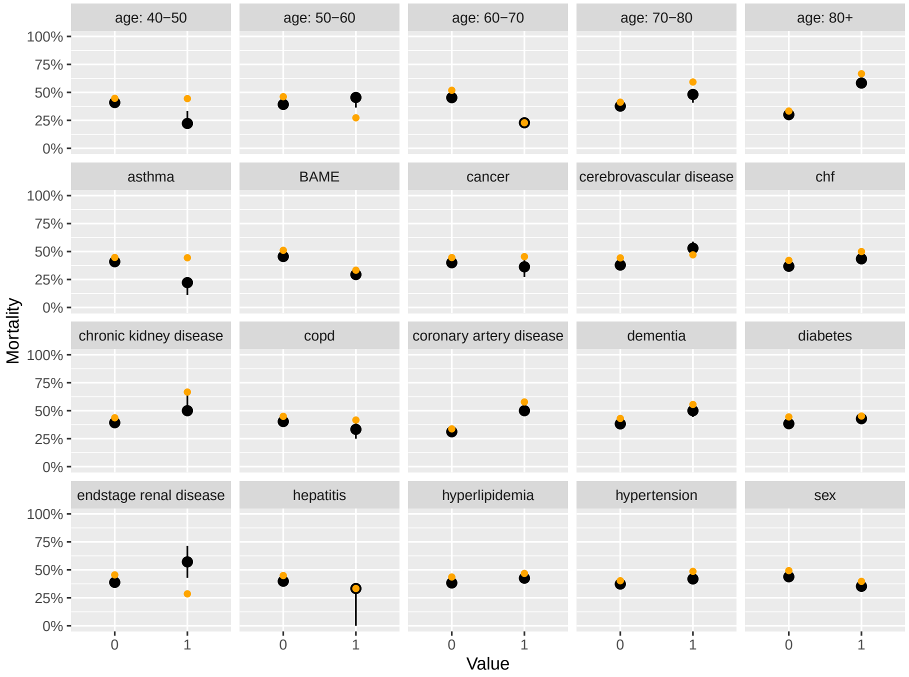


Figure S5: **Posterior predictive checks: demographic factors for Maimonides predicting an independent Maimonides dataset.** Each panel corresponds to a different binary variable, with the horizontal axis indicating its value. Orange points indicate observed mortality; black point-ranges indicate model estimated mortality, with the middle point indicating the posterior median, and the ranges indicating the 25%-75% ranges. The predictions were produced using the "post-admission" regression set in the Markov model.

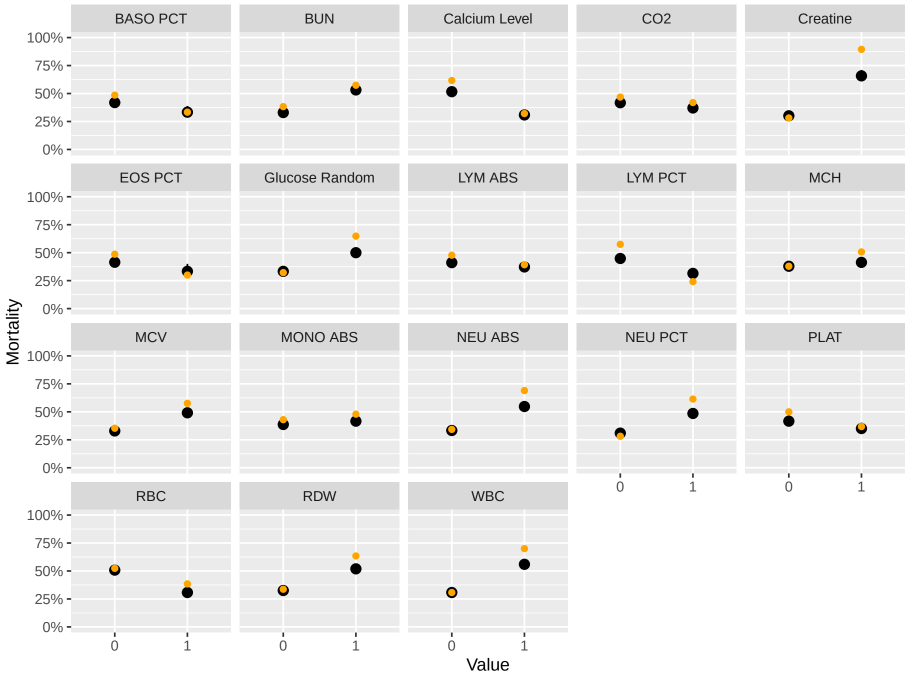


Figure S6: **Posterior predictive checks: dynamic factors for Maimonides predicting an independent Maimonides dataset.** Each panel corresponds to a different binarised variable (details of binarisation in text), with the horizontal axis indicating its value. Orange points indicate observed mortality; black point-ranges indicate model estimated mortality, with the middle point indicating the posterior median, and the ranges indicating the 25%-75% ranges. The predictions were produced using the "post-admission" regression set in the Markov model.

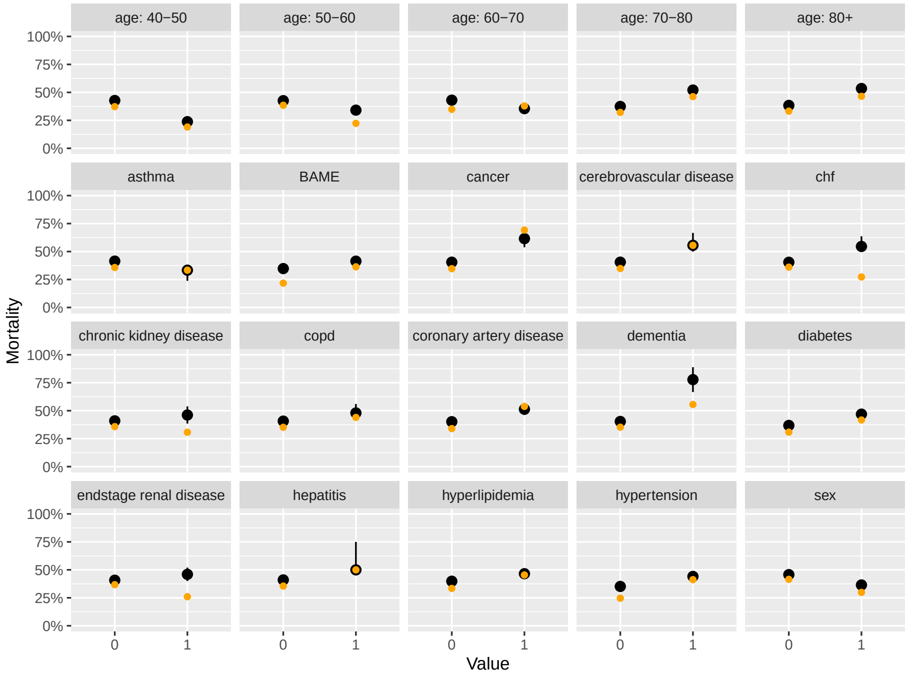


Figure S7: **Posterior predictive checks: demographic factors for Maimonides predicting an independent SUNY dataset.** Each panel corresponds to a different binary variable, with the horizontal axis indicating its value. Orange points indicate observed mortality; black point-ranges indicate model estimated mortality, with the middle point indicating the posterior median, and the ranges indicating the 25%-75% ranges. The predictions were produced using the "post-admission" regression set in the Markov model.

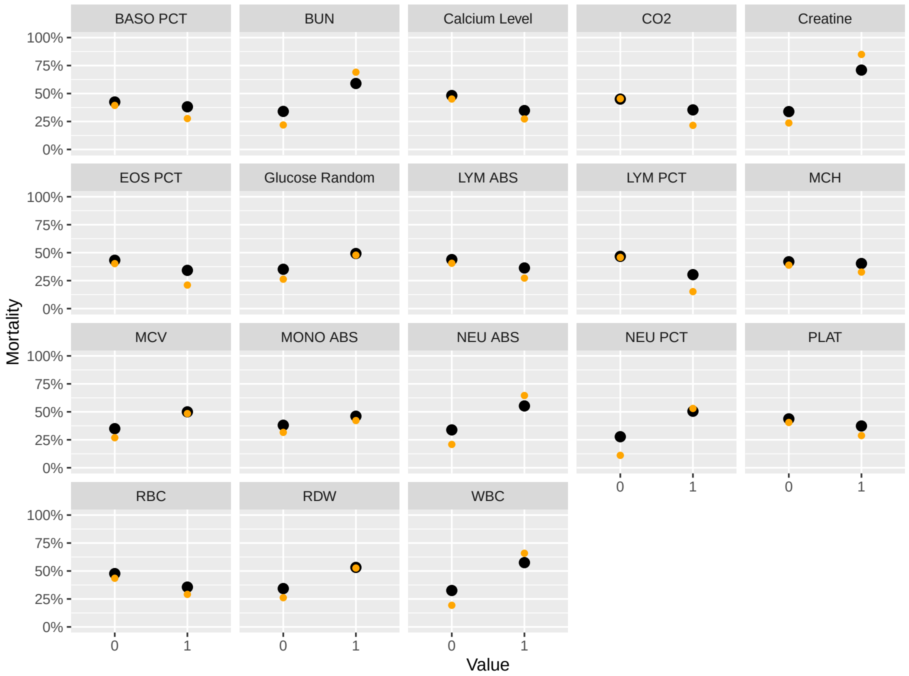


Figure S8: **Posterior predictive checks: dynamic factors for Maimonides predicting an independent SUNY dataset.** Each panel corresponds to a different binarised variable (details of binarisation in text), with the horizontal axis indicating its value. Orange points indicate observed mortality; black point-ranges indicate model estimated mortality, with the middle point indicating the posterior median, and the ranges indicating the 25%-75% ranges. The predictions were produced using the "post-admission" regression set in the Markov model.

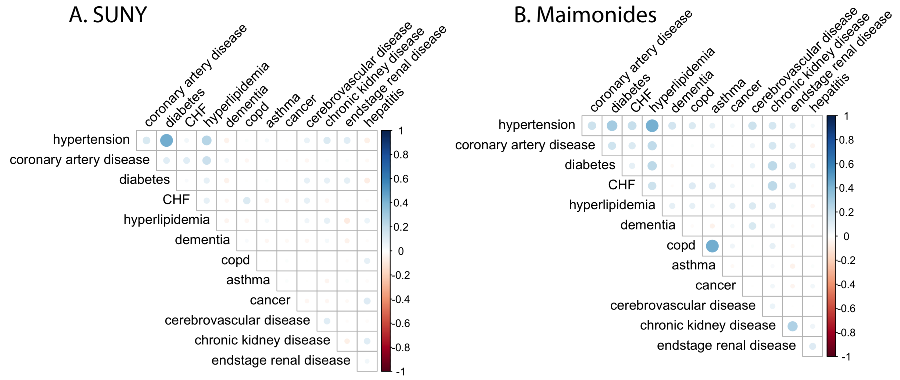


Figure S9: **Comorbidity correlations in each of the hospitals.** The color of each bubble indicates the sign and magnitude of the correlation in presence of paired correlations; the size of each bubble indicates the number of individuals with both conditions.

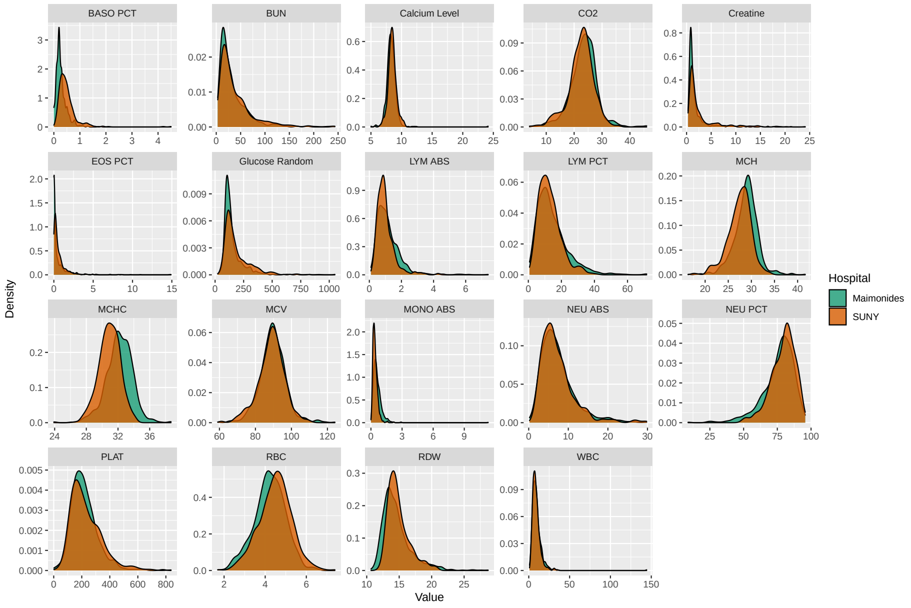


Figure S10: **Comparing laboratory values at admission across hospitals.** Each panel shows data for one of the $n = 19$ common tests across the hospitals.

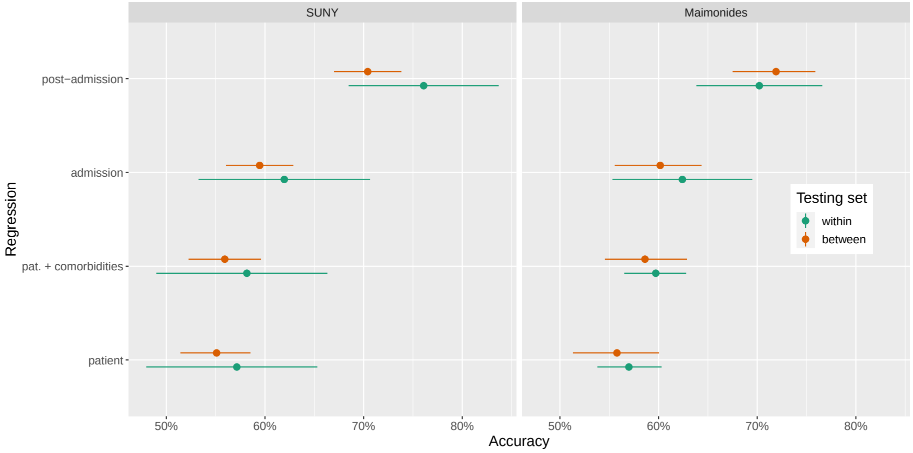


Figure S11: **Assessing between- versus within-hospital predictive accuracy.** The horizontal axis shows the accuracy in predicting patient outcomes (i.e. death or discharge) using a Markov regression model with covariate sets as named on the vertical axis. The points and whiskers indicate the posterior medians and 2.5%-97.5% posterior intervals for the percentage of patients whose outcome was correctly determined across MCMC draws. Each panel indicates the hospital whose data was used to train the model.

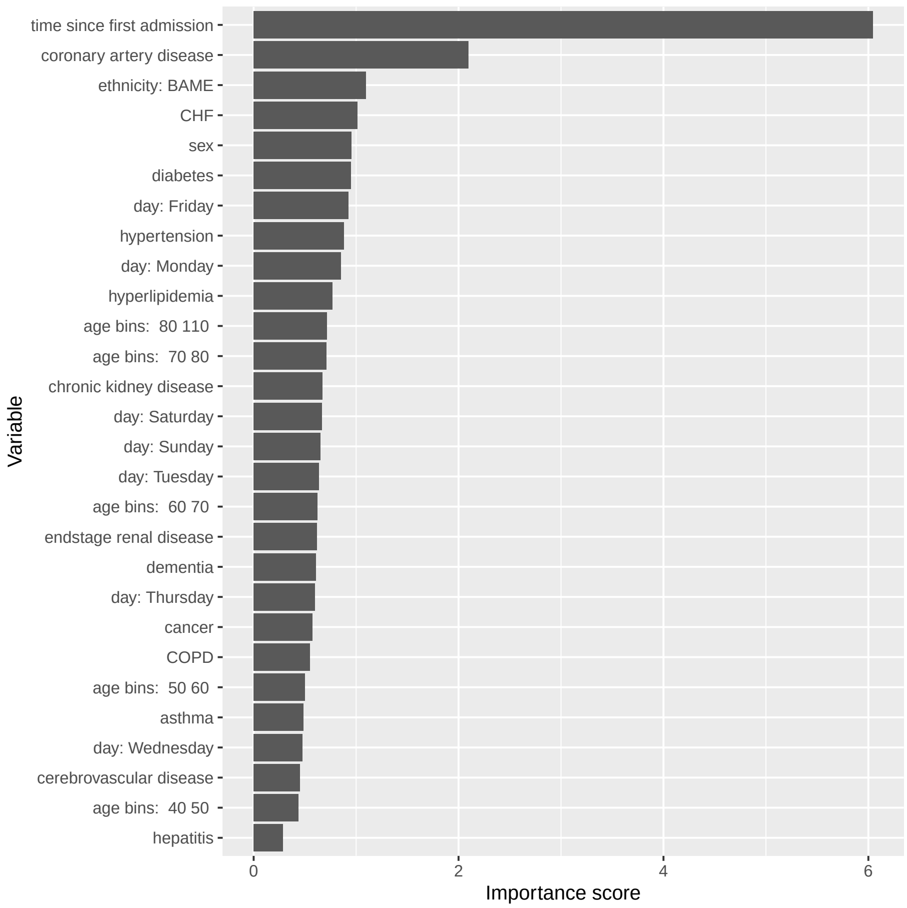


Figure S12: **Variable importance metrics from a Random Forest regression.** See text for more information.

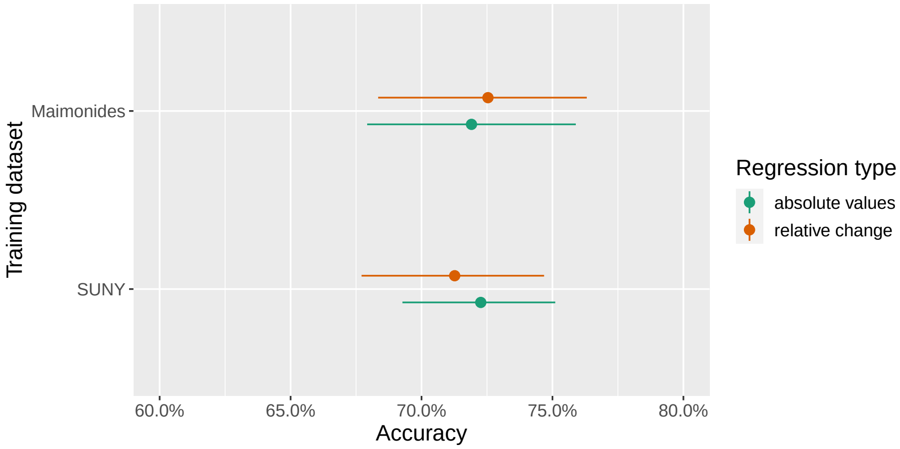


Figure S13: **Comparing the predictive power of raw biomarker values with the relative changes from baseline.** The horizontal axis shows the accuracy in predicting patient outcomes (i.e. death or discharge) using a Markov regression model across each hospital. The different colours indicate the two covariate types included in the analysis: either the raw biomarker values (denoted "absolute values") or the relative changes from baseline (denoted "relative changes"). The points and whiskers indicate the posterior medians and 2.5%-97.5% posterior intervals for the percentage of patients whose outcome was correctly determined across MCMC draws.

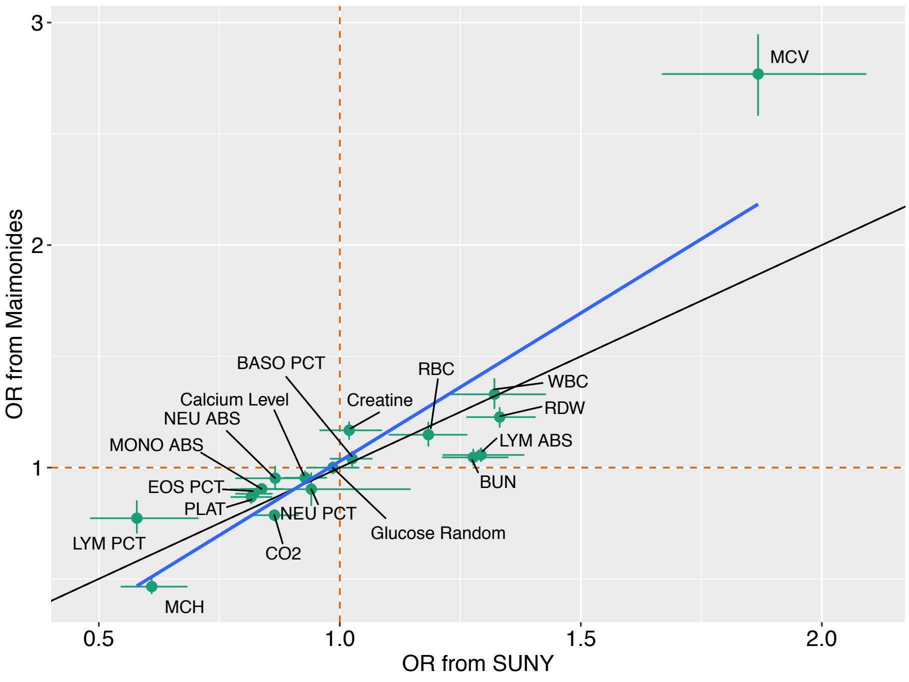


Figure S14: **Comparing the ORs for mortality risk for the dynamic factors across hospitals from the absolute values analysis.** The horizontal axes displays the ORs for daily mortality risk from SUNY and the vertical axes show the ORs from Maimonides. The ORs were estimated using the multivariate Markov model. Points show the posterior median ORs; the whiskers display the 25% and 75% posterior quantiles. The orange dashed lines show the OR = 1 cases; the dashed black lines indicate equality across ORs calculated across the hospitals. The blue line shows least squares regression lines using the posterior median ORs.

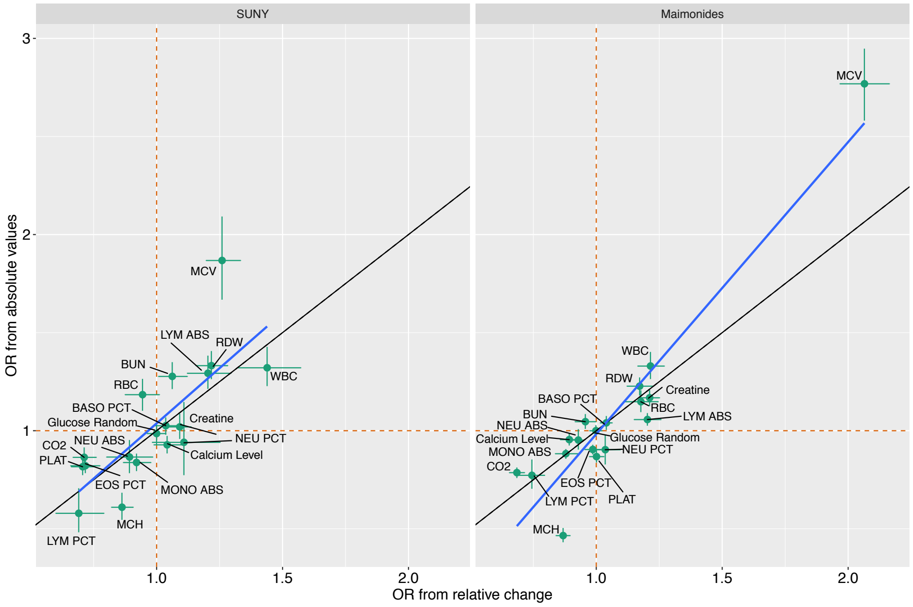


Figure S15: **Comparing the ORs for the relative changes analysis with those from the absolute values analysis.** The horizontal axes displays the ORs for daily mortality risk from the relative changes analysis and the vertical axes show the ORs from the absolute values analysis. The ORs were estimated using the multivariate Markov model. Points show the posterior median ORs; the whiskers display the 25% and 75% posterior quantiles. The orange dashed lines show the OR = 1 cases; the dashed black lines indicate equality across ORs calculated across the hospitals. The blue line shows least squares regression lines using the posterior median ORs. Each panel corresponds to a different hospital.
